# Association of mortality and aspirin use for COVID-19 residents at VA Community Living Center Nursing Homes

**DOI:** 10.1101/2022.08.03.22278392

**Authors:** Yasin Abul, Frank Devone, Thomas A Bayer, Christopher Halladay, Kevin McConeghy, Nadia Mujahid, Mriganka Singh, Ciera Leeder, Stefan Gravenstein, James L. Rudolph

## Abstract

**Background/Objectives:** Coronavirus disease 2019 (COVID-19) is associated with a hypercoagulable state and increased thrombotic risk in infected individuals. Several complex and varied coagulation abnormalities were proposed for this association^1^. Acetylsalicylic acid(ASA, aspirin) is known to have inflammatory, antithrombotic properties and its use was reported as having potency to reduce RNA synthesis and replication of some types of coronaviruses including human coronavirus-299E (CoV-229E) and Middle East Respiratory Syndrome (MERS)-CoV ^2,3^. We hypothesized that chronic low dose aspirin use may decrease COVID-19 mortality relative to ASA non-users.

**Methods:** This is a retrospective, observational cohort analysis of residents residing at Veterans Affairs Community Living Centers from December 13, 2020, to September 18, 2021, with a positive SARS-CoV-2 PCR test. Low dose aspirin users had low dose (81mg) therapy (10 of 14 days) prior to the positive COVID date and were compared to aspirin non-users (no ASA in prior 14 days). The primary outcome was mortality at 30 and 56 days post positive test and hospitalization.

**Results:** We identified 1.823 residents who had SARS-CoV-2 infection and 1,687 residents were eligible for the study. Aspirin use was independently associated with a reduced risk of 30 days of mortality (adjusted HR, 0.60, 95% CI, 0.40-0.90) and 56 days of mortality (adjusted HR, 0.67, 95% CI, 0.47-0.95)

**Conclusion:** Chronic low dose aspirin use for primary or secondary prevention of cardiovascular events is associated with lower COVID-19 mortality. Although additional randomized controlled trials are required to understand these associations and the potential implications more fully for improving care, aspirin remains a medication with known side effects and clinical practice should not change based on these findings.

## Introduction

To date, the severe acute respiratory syndrome coronavirus 2 (SARS-CoV-2) pandemic has affected over 1 million nursing home residents with a mortality rate over 13% ^4^ Nursing home residents have substantial risk for adverse outcomes due to age, frailty, and comorbidities. Thrombosis is a key feature of severe COVID-19; individuals with COVID-19 may have a variety of complex coagulation abnormalities that produce a hypercoagulable state. 5-30% of hospitalized COVID-19 patients suffer a major venous thromboembolic event, and up to 3% experience an arterial thromboembolic event, ischemic stroke, and myocardial infarction^5,6^. This brings into question appropriate assessments and interventions to prevent or treat thrombosis. Aspirin (ASA) may prevent severe COVID-19-related thrombotic effects through several mechanisms including inhibiting platelet activation and aggregation, decreasing platelet-driven inflammation, and preventing thrombogenic extracellular neutrophil rich environments. ASA can also affect viruses by blocking viral propagation, particularly in influenza and hepatitis C viruses ^7,8^. Additionally, ASA is known to play a beneficial role in the prevention and treatment of acute respiratory distress syndrome (ARDS), which can result from acute viral pneumonia, another major morbidity and mortality secondary to COVID-19 ^9^. The rise of breakthrough cases in people who are fully vaccinated is another cause of the continuing spread of SARS-CoV-2, with unknown outcomes particularly in the elderly. There is an emerging insight that pre-existing inflammation might be a factor of vaccine responsiveness. Controlling baseline inflammation before vaccination could be a promising solution to boosting vaccine responses ^10,11^. Multiple observational retrospective studies have shown that prior ASA use is associated with lower mortality rates in patients with COVID-19, albeit with discordant results from meta-analysis ^12-14^. On the other hand, one randomized controlled study of hospitalized patients with COVID-19 did not reveal ASA to have mortality benefit ^15^. There is no specific data particularly in frail nursing home patients about using aspirin in COVID-19 pandemics. The US Department of Veterans Affairs (VA) owns, operates, and manages nursing homes called Community Living Centers (CLCs). VA CLC residents are a unique population in which to study baseline chronic ASA use and its association with COVID-19 mortality.

## Aim of the Study and Hypothesis

Our study aim was to assess whether ASA use decreases mortality and hospitalization in people with COVID-19. We hypothesized that low dose ASA would be associated with decreased COVID-19 mortality and hospitalization.

## Study Methods and Design

### Description of Data

Data was taken from the VA Corporate Data Warehouse (CDW), a national, central database that collects and stores electronic health records from 1,255 VA health care facilities across the United States. The VA Health system has been linked on a common electronic medical record for 25 years. For this study, data on demographics, comorbidities, pharmacy, patient location, and vital status were linked from the VA Corporate Data Warehouse. The secondary analysis of clinical data was approved by the Providence VAMC IRB and R&D committees. Data release is governed by the VA Data Policy and is unable to be released at this time.

### Analytic Sample of the Study

This cohort study was conducted at VA CLCs. Among 19,759 CLC residents who had at least one COVID 19 test on record (Figure 1), 17,936 residents were excluded for their negative COVID-19 test status. 136 residents were excluded by intermittent/partial or high dose aspirin use. 1,687 VA CLC residents with SARS-CoV-2 infection confirmed by polymerase chain reaction testing between December 13, 2020, and September 18, 2021, were eligible for this study.

**Figure 1:**
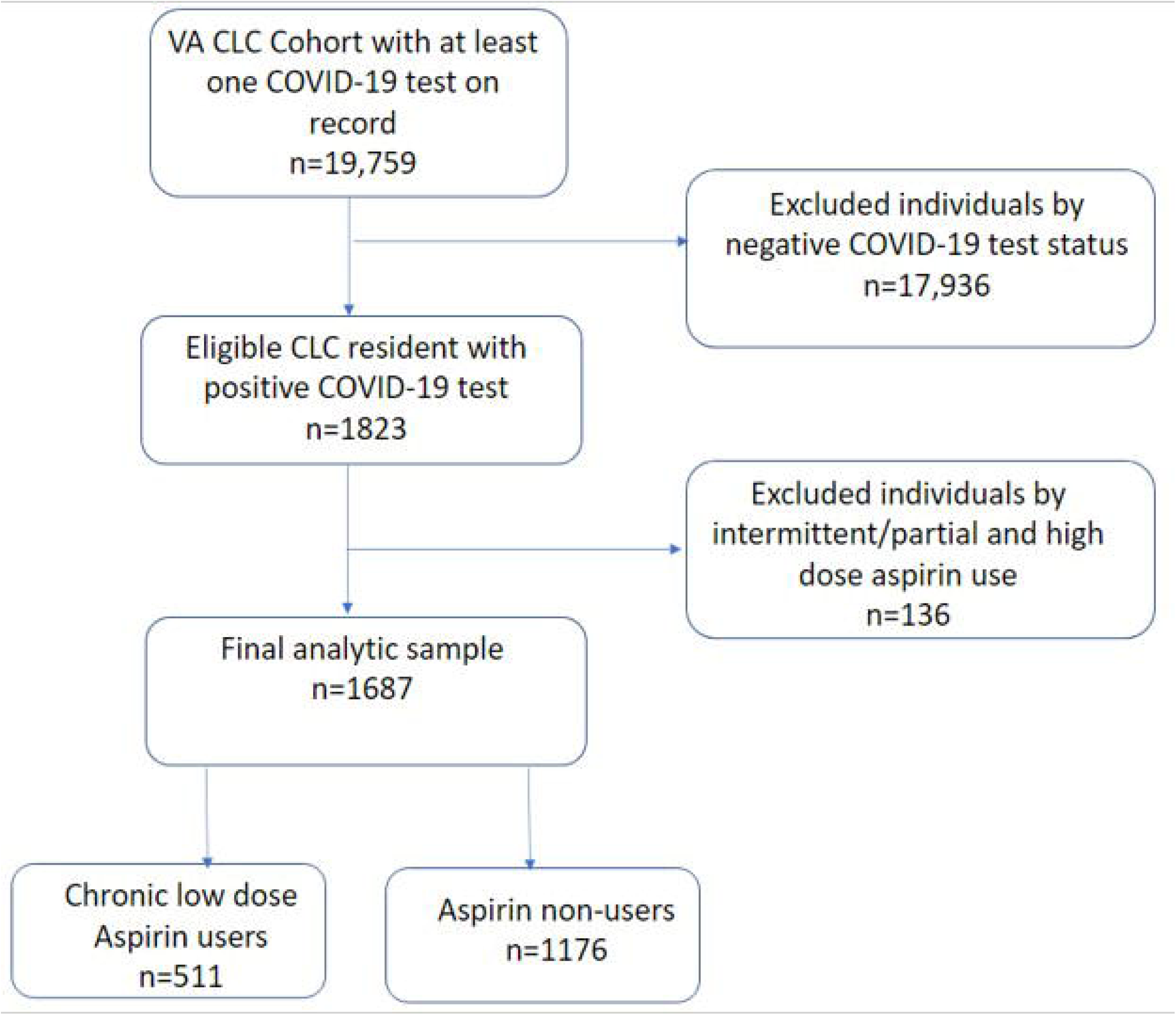
Consort diagram of CLC residents enrolled in the study

### Definitions of variables

#### Chronic low dose ASA user vs ASA non-user

Chronic low dose ASA users were identified as those taking low dose (81mg) ASA ten out of the 14 days before the date of the positive COVID-19 test result (index date). ASA non-users were defined as those who did not take any dose of ASA in the accrual window/exposure period (Figure 2). These definitions were chosen based on ASA’s prolonged duration of action as an irreversible platelet inhibitor which is approximately 10 days ^16-20^ High dose aspirin users and intermittent aspirin users were excluded to better define the homogenous chronic low dose aspirin user group (Figure 1)

**Figure 2:**
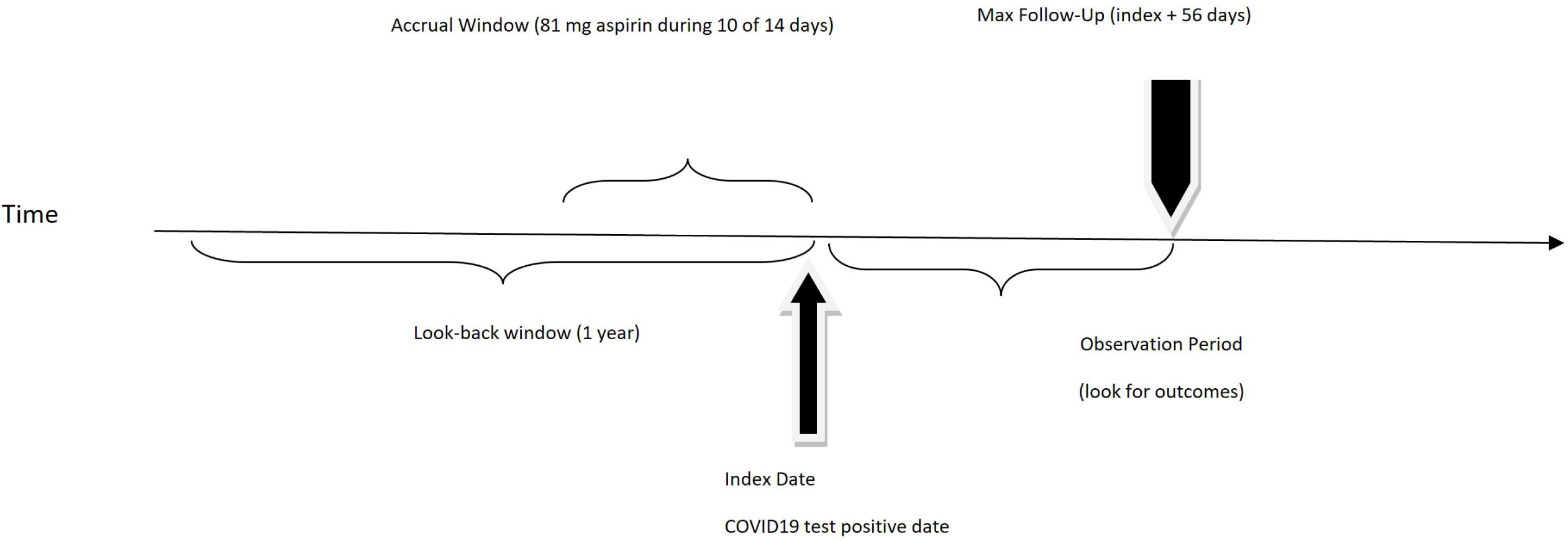
Recruitment timeline showing accrual window, look back window, observation window and maximum follow up days to visualize the study design

##### Exposure

The *c*onceptual exposure of interest was chronic low dose (81 mg) ASA use that was operationalized as use of aspirin 10 of 14 days prior to the positive COVID date.

##### Outcomes

The conceptual outcomes of interest in this study were mortality and hospitalization. Since death due to COVID-19 often occurs two to eight weeks after the onset of symptoms or a positive test, mortality was operationalized by calculating 30 days of mortality and 56 days of mortality ^21,22^. Hospitalization was defined as any resident hospitalized within 14 days of a laboratory-confirmed positive COVID-19 test according to CDC definition ^23^.

### Demographics and Covariates

The demographic characteristics of the data collected in the VA National CDW were chosen to summarize the study population. Gender was used as a binary variable with responses “male” and “female.” Race was classified as “white,” “African American,” and “other”. Potential major confounders/covariates of interest for this study included history of major adverse cardiac events (MACE), stroke, atrial fibrillation, myocardial infarction, heart failure, anemia, hematocrit level, and breakthrough infections.

### Analyses

Statistical analyses were conducted using R statistical software (Vienna, Austria Version 4.0.1). Standardized mean difference values farther from 0 indicate dissimilar groups, and values >0.1 can be interpreted as potentially meaningful differences. Cox proportional hazards models were used to define the adjusted associations between chronic low dose ASA use and outcomes including mortality and hospitalization after controlling for potential confounders.

## Results

From a total of 19,759 CLC residents with at least one COVID-19 test on record, we excluded residents with a negative COVID-19 test status (n=17,936) and those with SARS-CoV-2 infection who were high-dose, intermittent/partial aspirin users (n=136). The final analytic sample included 1,687 eligible residents with a mean age of 72.28±11.66 years and 3.3% (n=67) female. Compared to the ASA non-user group, the chronic low dose ASA user group had higher prevalence of history of stroke, diabetes, hypertension, major cardiac adverse events, congestive heart failure, and Alzheimer’s disease related dementias (Table 1).

**Table 1:** Baseline Characteristics of VA CLC residents by ASA use

|  | Overall | Low dose ASA User Group | ASA Non- User Group | SMD |
| --- | --- | --- | --- | --- |
| N | 1,687 (100%) | 511(30.3%) | 1,176(69.7%) | NA |
| Age | 72.28(11.6%) | 73.49 (9.5%) | 71.75(12.4%) | 0.156 |
| Male | 1,620(96.0%) | 494 (96.6%) | 1,126(95.7%) | 0.048 |
| Race: White | 1,145 (67.8%) | 357 (69.8%) | 788(67%) | 0.061 |
| Race: African American | 386 (22.8%) | 114 (22.3%) | 272(23.1%) | 0.019 |
| Race: Other | 156 (9.2%) | 40 (7.8%) | 116 (9.8%) | 0.071 |
| Breakthrough cases | 742 (4.9%) | 205 (40.1%) | 537 (45.6%) | 0.112 |
| Long stay (>100 days) | 970 (57.5%) | 354 (69.2%) | 616 (52.3%) | 0.351 |
| Previously hospitalized | 1,100(65.2%) | 289 (56.5%) | 811 (68.9%) | 0.258 |
| Prop days aspirin use | 0.30 (0.46) | 0.99 (0.04) | <10 (NA) | NA |
| Diabetes Mellitus | 846 (50.1%) | 318 (62.2%) | 528(44.9%) | 0.352 |
| Hypertension | 1,259(74.6%) | 404 (79.0%) | 855 (72.7%) | 0.149 |
| Congestive Heart Failure | 448 (26.5%) | 163 (31.9%) | 285(24.2%) | 0.171 |
| Chronic Pulmonary Diseases | 582 (34.5%) | 206 (40.3%) | 376(31.9%) | 0.174 |
| Valvular heart disease | 151(8.9%) | 45(8.8%) | 106(9.0%) | 0.007 |
| Alcohol use | 218 (12.9%) | 53(10.3%) | 165(14.0%) | 0.112 |
| Illicit drug use | 157(9.3%) | 38(7.4%) | 119(10.1%) | 0.094 |
| Anemia | 754 (44.6%) | 240 (46.9%) | 514 (43.7%) | 0.065 |
| Depression | 770 (45.6%) | 261 (51.0%) | 509 (43.2%) | 0.156 |
| Tumor | 295 (17.5%) | 73 (14.3%) | 222 (18.9%) | 0.123 |
| Psychoses | 487 (28.9%) | 166 (32.5%) | 321 (27.3%) | 0.113 |
| Renal disease | 389 (23.0%) | 138 (27.0%) | 251 (21.3%) | 0.133 |
| ADRD | 940 (55.7%) | 330(64.58%) | 610 (51.8%) | 0.259 |
| Atrial Fibrillation | 193 (11.4%) | 55 (10.7%) | 138 (11.7%) | 0.030 |
| TBI | 104 (6.1%) | 30 (5.8%) | 74 (6.29%) | 0.017 |
| MACE | 647 (38.3%) | 247 (48.3%) | 400 (34.0%) | 0.294 |
| Stroke | 416 (24.6%) | 195 (38.1%) | 221 (18.7%) | 0.439 |
| BUN >16.5 mg/dL | 860 (50.9%) | 291 (56.9%) | 569 (48.3%) | 0.172 |
| Sodium <135 mEq/L | 205 (12.1%) | 62 (12.1%) | 143 (12.1%) | 0.0008 |
| BNP > 100 pg/mL | 324 (19.2%) | 126 (24.6%) | 198 (16.8%) | 0.194 |
| Hematocrit Gendered Binary | 860 (50.9%) | 281 (54.9%) | 579 (49.2%) | 0.115 |
| Creatinine >1.5 mg/dL | 232 (13.7%) | 85 (16.6%) | 147 (12.5%) | 0.117 |
| One Pulse Ox reading <88% | 127 (7.5%) | 37 (7.2%) | 90 (7.6%) | 0.015 |

SMD: Standardized mean difference (values farther from 0 indicate dissimilar groups, and values >0.1 can be interpreted as potentially meaningful differences.). TBI: Traumatic brain injury; BNP: Brain natriuretic peptide; Pulse Ox: Pulse oximetry; ADRD: Alzheimer's disease related dementias; MACE: Major adverse cardiac events

On unadjusted analysis, the chronic low dose ASA user group, compared to the ASA non-user group had significantly lower mortality rates: 30 days (6.46% [33/511] for ASA user group versus 10.29% [121/1176] for ASA non-user group, SMD >0.1), and 56 days (9.0% [46/511] ASA user group versus 13.18% [155/1176] ASA non-user, SMD >0.1) (Table 1).

After adjusting for confounding variables in the Cox proportional hazards model, aspirin use was independently associated with a reduced 30 day and 56-day mortality risk (adjusted HR, 0.60, 95% CI, 0.40-0.90 for 30 days of mortality; adjusted HR, 0.67, 95% CI, 0.47-0.95 for 56 days of mortality) (Table 2, Figure 3).

**Table 2:** Cox Proportional Hazard model to examine the effect of ASA on mortality

| Variable | Low Dose ASA user group | ASA non-user group | SMD | Unadjusted HR (95% of CI) | Adjusted HR (95% of CI) |
| --- | --- | --- | --- | --- | --- |
| 30 days of mortality | 154 (9.1%) | 33 (6.4%) | 121 (10.2%) | 0.61 (0.41-0.89) | 0.60 (0.4- 0.9) |
| 56 days of mortality | 201 (11.9%) | 46 (9%) | 155 (13.1%) | 0.66 (0.47-0.92) | 0.67 (0.47-0.95) |
| Hospitalization | 352 (20.8%) | 103 (20.1%) | 249 (21.1%) | 0.88 (0.67-1.14) | 0.8 (0.60-1.07) |

SMD: Standardized mean difference (values farther from 0 indicate dissimilar groups, and values >0.1 can be interpreted as potentially meaningful differences.). HR: Hazard ratio; CI: Confidence interval

**Figure 3:**
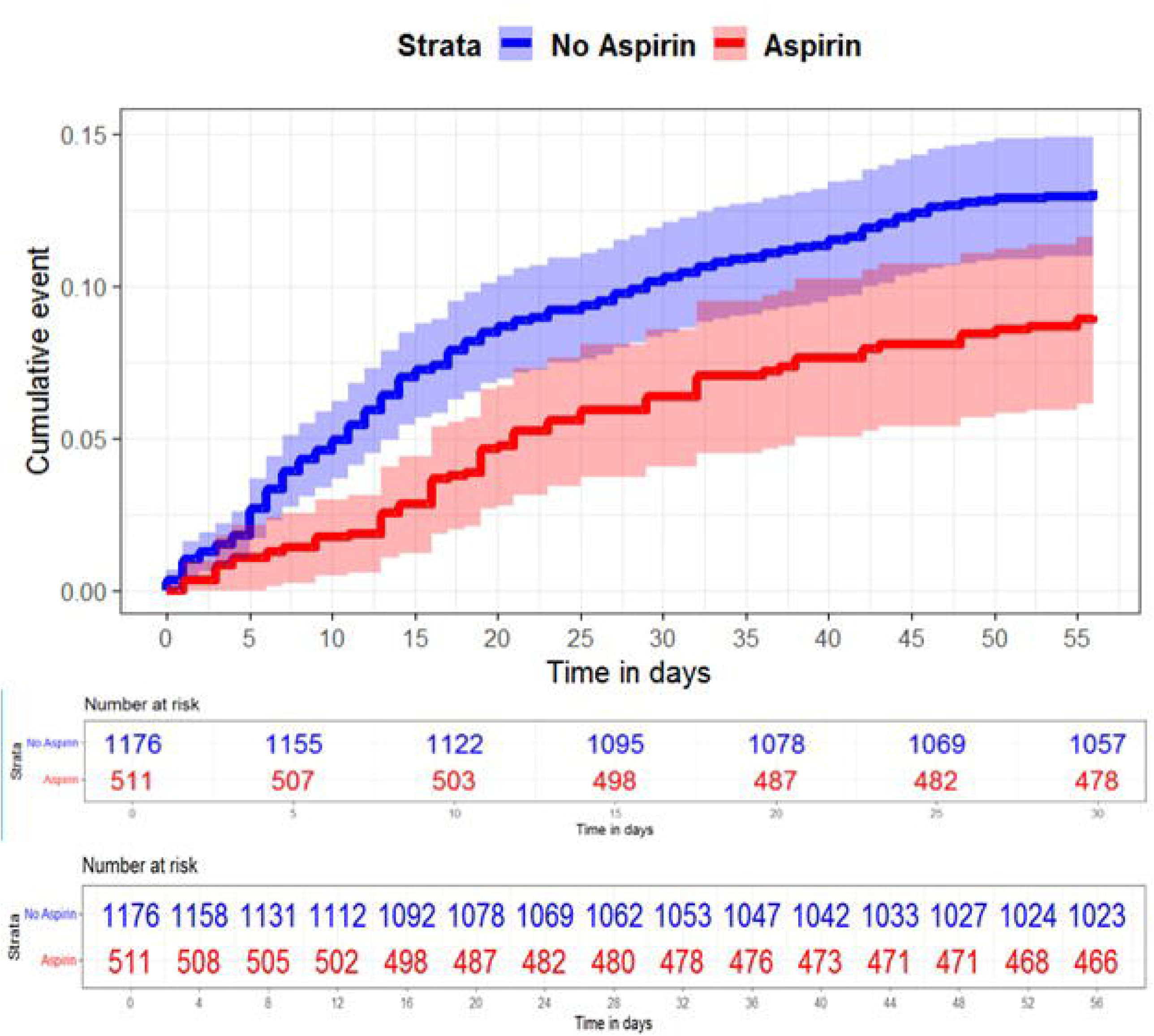
Longitudinal risk outcome in those with/without exposure, survival curve for 30 and 56 days of mortality. Patients are stratified by aspirin use. Patients that reached the maximum follow-up observation period are right-censored. ASA use was associated with a decreased hazard for 30 and 56 days of mortality (adjusted HR, 0.60, 95% CI, 0.40-0.90 for 30 days of mortality; adjusted HR, 0.67, 95% CI, 0.47-0.95 for 56 days of mortality). CI indicates confidence interval; HR indicates hazard ratio.

## Discussion

This study showed that there is an association between chronic low dose use of ASA and a lower mortality rate in CLC residents with COVID-19, compared with non-ASA users. Time-to-event analysis showed the survival difference between the chronic low dose ASA user group and ASA non-user group with the mortality rate favoring the chronic low dose ASA user group after adjusting for other covariates.

Literature supports that ASA may reduce mortality in hospitalized patients with COVID, but there is less evidence in nursing home residents with COVID. Prior work on the association between aspirin use and outcomes of COVID present conflicting findings. Observational retrospective cohort data showed that baseline ASA use was associated with lower mortality compared with those who did not use ASA, particularly in hospitalized patients. This study showed the association of chronic low dose ASA use and lower mortality rate in CLC residents. The meta-analyses reported different conclusions about the effect of ASA on mortality in COVID-19 ^12-14^.

First meta-analysis included 3 studies and showed mortality among aspirin users was 22.6% with respect to mortality of 18.3% among non-aspirin users (risk ratio 1.12, 95% confidence intervals [0.84, 1.50]) ^12^. Second meta-analysis included six eligible studies and indicated the use of low-dose aspirin was independently associated with a reduced mortality (RR 0.46 [95% confidence interval 0.35–0.61], P < 0.001; I² = 36.2%) ^14^. Third and largest meta-analysis included seven studies and concluded that the use of aspirin was associated with a reduced risk of mortality (RR 0.56, 95% CI 0.38–0.81, P = 0.002; I²: 68%, P = 0.005) ^13^.

ASA use was reported previously as independently associated with decreased risk of hospital mortality, mechanical ventilation, and intensive care unit (ICU) admission ^24^. A prior ASA prescription was associated with decrease in overall mortality at 14 days and 30 days in COVID-19-positive veterans as well ^25^. On the other hand, one randomized, controlled, open-label, platform trial showed that ASA was not associated with decreased 28-day mortality rate, risk of disease progression leading to the need for invasive mechanical ventilation, or death in patients hospitalized with COVID-19. However, it did show that ASA was associated with a small surge in live hospital discharge within 28 days. Additionally, a small reduction in thrombosis was associated with ASA use (4.6% vs 5.3%) and minimal increase in major bleeding (1.6% vs 1.0% ^15^. None of the studies particularly investigate and emphasize possible effect of ASA use before the index date, especially before or during the 5 days incubation period of COVID-19 which could be actual reasoning of different results of meta-analyses.

Previous observational cohort studies have included dosages of ASA ranging from 75-150mg (75mg, 81mg,100mg, and 150mg respectively) and administered at various times in the clinical course of COVID-19 ^13,24-29^. In contrast to previous observational studies of the effects of aspirin in COVID-19, our precise exposure definition assures consistency of treatment. Our inclusion of only long-stay CLC residents makes our findings especially relevant to the care of long-term care residents, a medically complex population who has suffered a disproportionate burden of COVID-19 morbidity and mortality.

The association between ASA use and lower mortality in CLC residents with COVID-19 may be related to multiple factors. First, the antithrombotic effect of ASA could have an impact on prothrombotic, and hypercoagulable states induced by COVID-19. Through this mechanism, ASA could be proposed to prevent thrombotic complications including COVID-related myocardial infarction, stroke, deep vein thrombosis, arterial thrombosis, or pulmonary embolism ^30-33^. Second, the proposed protective effect might be from the antiviral properties of ASA which has previously been shown for viruses of H1N1, MERS-CoV, and CoV-229E ^3,13,34^. This antiviral property is modulated by different pathways including the nuclear factor kappa beta (NF-κB) pathway ^3,35^.

It is acceptable to keep low dose aspirin therapy if the individual is already on it for another indication and to use low dose aspirin for standard indications. However, contraindications and adverse effects of aspirin must be always in consideration, particularly in relation to bleeding risk in frail, elderly population. Given the low certainty of current data, the risk benefit ratio should be assessed before starting aspirin in individuals with COVID-19.

## Strengths and Limitations

The present study limits generalizability due to its retrospective observational design, and the study population’s small sample size, male predominance, and higher frequency of multiple comorbidities. Another limitation is the presence of potential diverse, unpredictable, and unmeasured confounders.

Next steps would be to perform further sensitivity analysis to understand if the association between ASA use and mortality is a true cause and effect association and if there are other confounding factors such as concomitant use of other antithrombotic medication use.

## Conclusion

Low dose ASA use is associated with lower mortality in VA CLC residents with COVID-19. While the association is important to explore further, ASA remains a medication with known side effects and clinical practice should not change based on these findings, particularly in long-term, frail nursing home residents. It is acceptable to use ASA for standard indications and to continue ASA if the patient is on it for different evidence-based indications.

## Data Availability

The secondary analysis of clinical data was approved by the Providence VAMC IRB and R&D committees. Data release is governed by the VA Data Policy and is unable to be released at this time.

## Acknowledgement

The authors thank Margo Katz for providing editorial assistance for this manuscript

